# Mass-Standardised Differential Antibody Binding to a Spectrum of SARS-CoV-2 Variant Spike Proteins: Wuhan, Alpha, Beta, Gamma, Delta, Omicron BA.1, BA.4/5, BA.2.75 and BA.2.12.1 variants - Antibody Immunity Endotypes

**DOI:** 10.1101/2022.09.23.22280271

**Authors:** Philip H. James-Pemberton, Shival Kohli, Jordan Twynham, Aaron C. Westlake, Alex Antill, Rouslan V. Olkhov, Andrew M. Shaw

**Author notes:** **Declaration of Interest** Prof Shaw is the Founder, CEO and Director of Attomarker Ltd, a spin-out company from his research group. **Related Publications** **Paper 1** - Mass-Standardised Quantitative Measurements of the Antibody Levels for SARS-CoV-2 beyond Correlates of Protection and Clearance **Paper 2** – This Paper - Mass-Standardised Differential Antibody Binding to a Spectrum of SARS-CoV-2 Variant Spike Proteins: Wuhan, Alpha, Beta, Gamma, Delta, Omicron BA.1, BA.4/5, BA.2.75 and BA.2.12.1 variants - Antibody Immunity Endotypes **Paper 3** –Mass-Standardised Antibody Affinity Maturation to the Spike Protein of SARS-CoV-2 Omicron Variants in a Teachers Cohort: forgiving Original Antigenic Sin **Paper 4** – Diagnostic Test for Long Covid Patients pointing to a Classification of Persistent Virus and Autoimmune Pathophysiologies leading to a Test-and-Treat Protocol.

## Abstract

A fully mass-standardised quantitative comparative analysis of the differential binding to spike variant proteins to SARS-CoV-2 has been performed for the variants: Wuhan, Alpha, Beta, Gamma, Delta, and the Omicron variants BA.1, BA.2.12.1, BA.2.75, BA.4 and BA.5. Evolution of immunity through five patient cohorts (*n* = 148 in total) was studied including pre-pandemic, first infection, first vaccine, second vaccine and triple-vaccinated cohorts. A population of immunity endotypes has been observed and is classified against a recovery antibody threshold. U(+) showing protection to all variants; single, double, triple, and further dropout endotypes U(±); some with no variant protection other than Wuhan vaccine spike U(−); and some unclassified, U(∼). These endotypes may be imprinted. In the triple-vaccinated cohort (*n* = 41) there is a U(+) incidence of 54% (95% CI 39% - 68%) suggesting between half and three-quarters of the population have universal variant vaccine antibody protection; and U(±)) with at least one dropout has an incidence of 42% (95% CI 28% - 57%). Extending the cohort incidence to the population, up to 68% of the population may have an imprinted immunity endotype to an epitope that is effective against all variants; critical for both protection and binding to the ACE2 receptor: a universal immunity endotype: up to 13% may not have a sterilising serum leading persistent virus and a risk of long covid.

## Introduction

The response to immunisation is personalised with antibody production based on affinity-matured clones selected against epitopes on the spike proteins of the SARS-CoV-2 pathogen^1^. Vaccination and infection, ideally, should mature antibodies and T cells to conserved epitopes that are resistant to mutation. However, the choice of the affinity-matured epitope may depend on the first exposure to the antigen which is then retained in B cells and produced for all subsequent exposures to the antigen – the concept of original antigenic sin^2–4^. If the epitope is not conserved during mutation of the viral protein target, the antibody response may become significantly impaired. The rapid mutations observed^5^ in SARS-CoV-2 made the initial vaccines redundant, well-targeted to the Wuhan variant but quickly evaded by later variants. Bivalent mRNA vaccines produce spike proteins for both the Wuhan and an Omicron variants, BA.5^6^ (Pfizer) or BA.1^7^ (Moderna) and most recently FDA approval of vaccines targeting the KP.3 strain. Some studies with different vaccine combinations^8,9^ have shown improvement in the resulting neutralising antibody protection.

Evaluation of vaccine efficacy needs to keep pace with the evolution of the virus and hence harmonised^10^ correlates of protection (CoP) and correlates of clearance (CoC) are required to bridge the variants or be re-derived. Ideally, the CoPs and CoCs should be replaced with a mass-standardised mechanism of protection and clearance which are not dependent on the variants, that point to accurate characterisation of the serum antibody profile^11^. By comparison, CoPs are variant specific^12^. Five-fold variation Receptor Binding Domain (RBD)-ACE2 binding affinities have been observed for different variants^13^; five-fold RBD-ACE2 association and dissociation rates^14^ also show variant dependence; and more interestingly increased viral load in later variants^15^. Neutralising antibody (NAb) assays have shown decline in antibody titres with variant^16^ reflecting the conserved epitope map in the NAbs produced in the patient response triggered by vaccine, infection or complex exposure pattern.

In this paper, we explore the variation of the Mass-standardisation (NIST standard antibody^11^) of individual patient response to the set of SARS-CoV-2 variants: Wuhan, Alpha, Beta, Gamma, Delta and Omicron BA.1, BA.4, BA.5, BA.2.75 and BA.2.12.1. Comparison with the mechanistic threshold derived previously^11^ of 1.8 mg/L (0.2 – 3.4 mg/L (95% CI)) was used to assess patient immunotype. Patient samples were screened from five cohorts with varying infection and vaccine profiles (*n* = 148 in total); pre-pandemic (controls); the Wuhan wave pre-vaccine; and then single, fully vaccinated and boosted cohorts for select combinations of vaccines, boosters and infections. A classification of immunity endotype is derived from the observation of different immune responses for protection and clearance.

## Methods and Materials

### Methods

#### Biophotonic Multiplexed Immuno-kinetic assay

A biophotonic platform, such as localised particle plasmon or continuous gold sensor, fundamentally consists of effective-mass sensors responding to the changes in refractive index in the plasmon field once it has been excited. The localised particle plasmon technique used here utilises an immuno-kinetic assay which has been described in detail elsewhere in a SARS-CoV-2 antibody sensing application^11,17^. Briefly, gold nanoparticles are printed into an array of 170 spots which are then individually functionalised with the SARS-CoV-2 spike proteins from each of the variants described in Table S1, alongside control spots of recombinant Human Serum Albumin for variation in illumination, temperature and non-specific binding. A protein A/G control channel measures total IgG and is used to assess antibody integrity in the calibration reagents.

The integrity of the protein samples on the surface was tested using a panel of antibodies raised to RBD, S1, S2 or whole S protein, detailed in Table S2. An anti-S2 antibody (40590-D001) was chosen for calibration as the corresponding epitope is present on all variants. The concentration of the antibody was calibrated against the NIST antibody RM8671, NISTmAb, a recombinant humanized IgG1ĸ with a known sequence^18^ to assure monomeric purity of the antibody. High-control and low-control samples were made using the 40590-D001 antibody and these controls were then used to quantify results from human samples in units of mg/L.

All 96 samples were collected within one antibody one half-life (60 – 200 days)^19^, 23 samples were collected above 60 days, Table 1. The median days since the double-vaccination was 43 days (26 – 121 days lower and upper quartile) and for the triple-vaccinated cohort the median was 32 days (20 – 63 days lower and upper quartile). A technique-independent endotype classification is based on the diagnostic accuracy threshold derived from four patient cohorts: recovering from infection (*n* = 200 PCR(+) and 200 controls); percentile of two vaccine response distributions that are resistant to live viral challenge; and a set of circuiting IgG positive standards^11^. The average of all thresholds is 1.8 mg/L (95% CI 0.2 – 3.4 mg/L) and a universal positive endotype, U(+), has the antibody concentrations for all variants above the 1.8mg/L threshold. A single drop-out has an antibody concentration for one variant below the threshold, *e.g.* β(−), double drop-out, two variants α(−)β(−) *etc*., for all variants and these are all labelled U(±); one or more variant antibody concentrations <1.8 mg/L. U(−) has a dropout below threshold for all variants. A further classification of transient endotypes, UT, was made to allow for waning of antibody concentration with time; the antibody levels were doubled and UT(+), UT(±) and UT(−). Wilson 95% confidence limits were calculated for the proportions of each endotype in the cohorts.

**Table 1.**
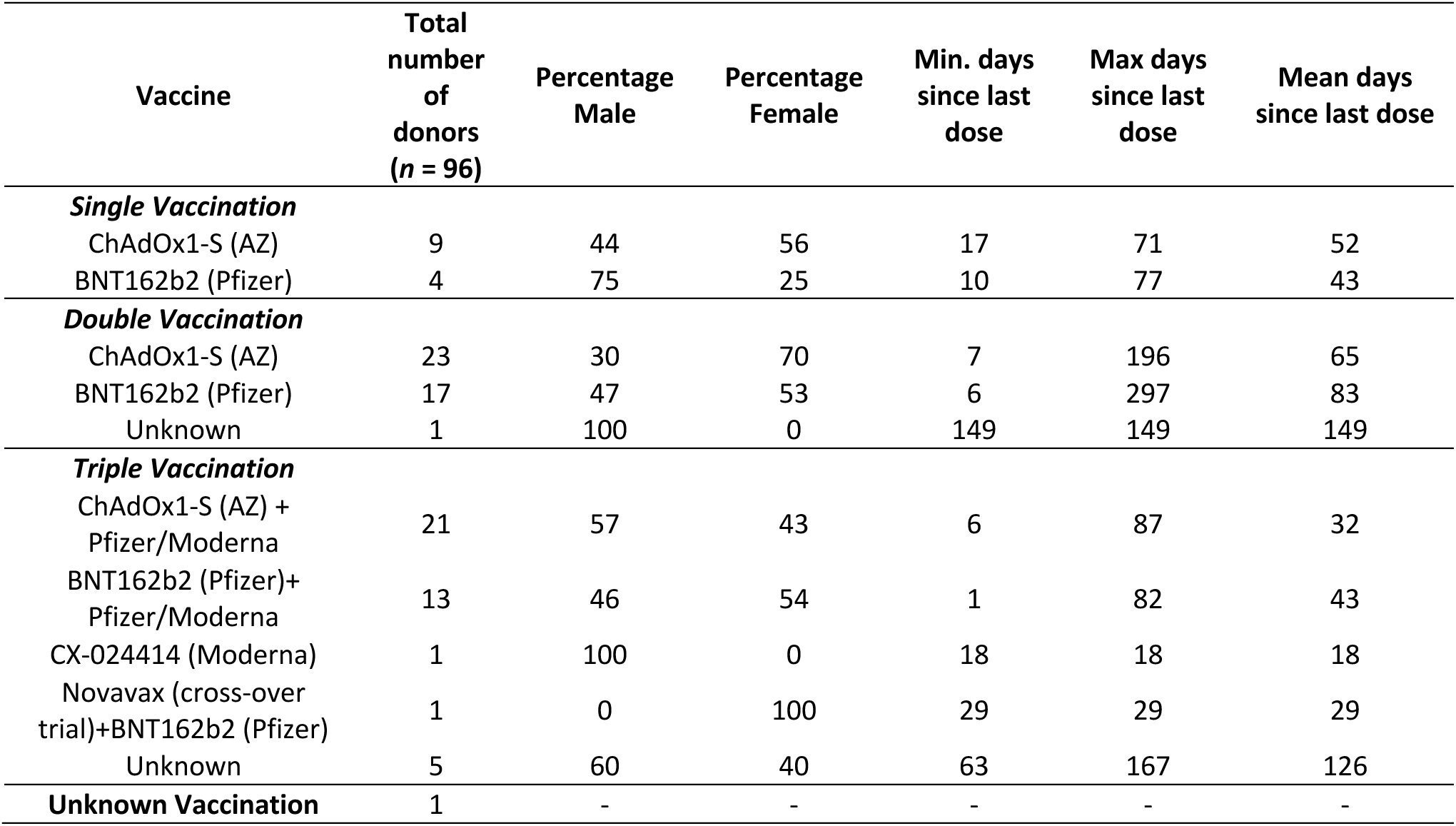
Demographic data for samples from vaccinated individuals collected in the Attomarker clinic.

| Vaccine | Total number of donors (n = 96) | Percentage Male | Percentage Female | Min. days since last dose | Max days since last dose | Mean days since last dose |
| --- | --- | --- | --- | --- | --- | --- |
| **Single Vaccination** |  |  |  |  |  |  |
| ChAdOx1-S (AZ) | 9 | 44 | 56 | 17 | 71 | 52 |
| BNT162b2 (Pfizer) | 4 | 75 | 25 | 10 | 77 | 43 |
| **Double Vaccination** |  |  |  |  |  |  |
| ChAdOx1-S (AZ) | 23 | 30 | 70 | 7 | 196 | 65 |
| BNT162b2 (Pfizer) | 17 | 47 | 53 | 6 | 297 | 83 |
| Unknown | 1 | 100 | 0 | 149 | 149 | 149 |
| **Triple Vaccination** |  |  |  |  |  |  |
| ChAdOx1-S (AZ) + Pfizer/Moderna | 21 | 57 | 43 | 6 | 87 | 32 |
| BNT162b2 (Pfizer)+ Pfizer/Moderna | 13 | 46 | 54 | 1 | 82 | 43 |
| CX-024414 (Moderna) | 1 | 100 | 0 | 18 | 18 | 18 |
| Novavax (cross-over trial)+BNT162b2 (Pfizer) | 1 | 0 | 100 | 29 | 29 | 29 |
| Unknown | 5 | 60 | 40 | 63 | 167 | 126 |
| **Unknown Vaccination** | 1 | - | - | - | - | - |

A full correlation analysis for all combinations of responses was also performed to assess whether responses to one variant correlated with or predicated the response to another variant: correlation coefficients and lines of best fit were derived.

### Materials

Materials used throughout the course of the experiments were used as supplied by the manufacturer, without further purification. Sigma-Aldrich supplied phosphate buffered saline (PBS) in tablet form (Sigma, P4417), phosphoric acid solution (85 ± 1 wt. % in water, Sigma, 345245) and Tween 20 (Sigma, P1379). Glycine (analytical grade, G/0800/48) was provided by Fisher Scientific. Assay running and dilution buffer was PBS with 0.005 v/v % Tween 20 and the regeneration buffer was 0.1 M phosphoric acid with 0.02 M glycine solution in deionized water.

The recombinant human antibody to the spike protein S2 subdomain was a chimeric monoclonal antibody (SinoBiological, 40590-D001, Lot HA14AP2901). The antibody was raised against recombinant SARS-CoV-2 / 2019-nCoV Spike S2 ECD protein (SinoBiological, 40590-V08B). The panel of antibodies seen in Table S2 were used to screen antigens for relative integrity and epitope presentation.

NISTmAb, Humanized IgG1κ Monoclonal Antibody from National Institute of Standards and Technology (RM8671). The NISTmAb is a recombinant humanized IgG1ĸ with a known sequence^18^ specific to the respiratory syncytial virus protein F (RSVF)^20^. The detection mixture consisted of a 200-fold dilution of IG8044 R2 from Randox in assay running buffer.

Two sensor chips designs were printed; an Omicron focussed array with recombinant human serum albumin (rHSA) from Sigma-Aldrich (A9731), protein A/G (PAG) from ThermoFisher (21186), and six SARS-CoV-2 Spike Protein variants for the Wuhan and trimer proteins for Omicron BA.1, BA.2.12.1, BA.4, BA.5 strains from SinoBiological and BA.2.75 spike protein from Acro Biosystems. The other sensor chip design included earlier variants, with rHSA, PAG and six monomer Spike Protein variants: Wuhan, Alpha, Beta, Gamma, Delta and Omicron BA.1. The complete protein data can be found in Table S1.

### Patient Samples

#### Commercial samples

Serum samples was purchased from two suppliers (Biomex GmbH and AbBaltis). 17 pre-pandemic (pre-December 2019), PCR(−) human serum samples were purchased from AbBaltis. All were tested and found negative for STS, HbsAg, HIV1 Ag (or HIV PCR(NAT)), HIV1/2 antibody, HCV antibody and HCV PCR(NAT) by FDA approved tests. 11 positive samples were purchased from AbBaltis which were all from PCR(+) individuals. No information was provided regarding symptoms of the donors. 45% of these samples were from female donors and 55% were from male donors. The age of donors ranged from 19 to 81 years. No information on time from infection to sample collection was given.

Samples purchased from Biomex (*n*=26) were from PCR(+) individuals. All samples were YHLO Biotech SARS-CoV-2 IgG positive and Abbott SARS-CoV-2 IgG positive. A spectrum of patient symptoms from the following list were detailed for each sample: fever, limb pain, muscle pain, headache, shivers, catarrh, anosmia, ache when swallowing, diarrhoea, breathing difficulties, coughing, tiredness, sinusitis, pneumonia, sickness, lymph node swelling, pressure on chest, flu-like symptoms, blood circulation problems, sweating, dizziness, and hospitalisation. Time from infection to sample collection ranged from 27 to 91 days. 35% of these samples were from female donors and 65% were from male donors. The samples were collected prior to June 2020; PCR(+) samples will result from infection by the SARS-CoV-2 strains circulating prior to this time, most similar to the Wuhan protein. These commercial samples, 38(+) and 16(−), were tested using the Spike Variant Array as a part of this study.

#### Clinical samples

Samples were collected from patients in Attomarker partner clinics, all of whom provided informed consent for their anonymised data to be used in research to aid the pandemic response. The data from tests of 96 patient samples are included in this study: 13 had received one dose of either AstraZeneca (AZ) or Pfizer SARS-CoV-2 vaccine; 41 had received two doses of either AZ, Pfizer or Moderna SARS-CoV-2 vaccine at least 14 days prior to sample collection and testing by Attomarker; 41 further samples were from patients who had received a third vaccination. Full patient demographics are shown in Table 1. One sample had no vaccine history disclosed.

#### Ethical Approval

The use of the Attomarker clinical samples with the consent of the patients was approved by the Bioscience Research Ethics Committee, University of Exeter

## Results

The pan-variant integrity of a chimeric monoclonal antibody to the S2 region of the Spike protein was established by measuring the antibody binding to the spike protein panel, deriving a binding maximum within a Langmuir adsorption model (Table S3) as a quantitative measure of site density. The antibody was then used as the calibrant for the antibody binding spectrum, mass-standardised against the NIST standard antibody. The antibody binding variant spectra and endotype classification of some patient responses are shown in Figure 1, with the population distributions for all samples shown in Figure 2. The combined patient cohorts showed a range of immunity endotypes in response to the variant spectrum of SARS-CoV-2 spike proteins. The pre-pandemic, negative control samples were U(−) = 81% (95 CI 57% - 93%) spike antibody negative with 3 patients showing 9-fold dropouts suggesting immunity potentially due to a misclassification: a PCR false-positive, previous alphacoronavirus infection or random antibody cross-reactivity.

**Figure 1.**
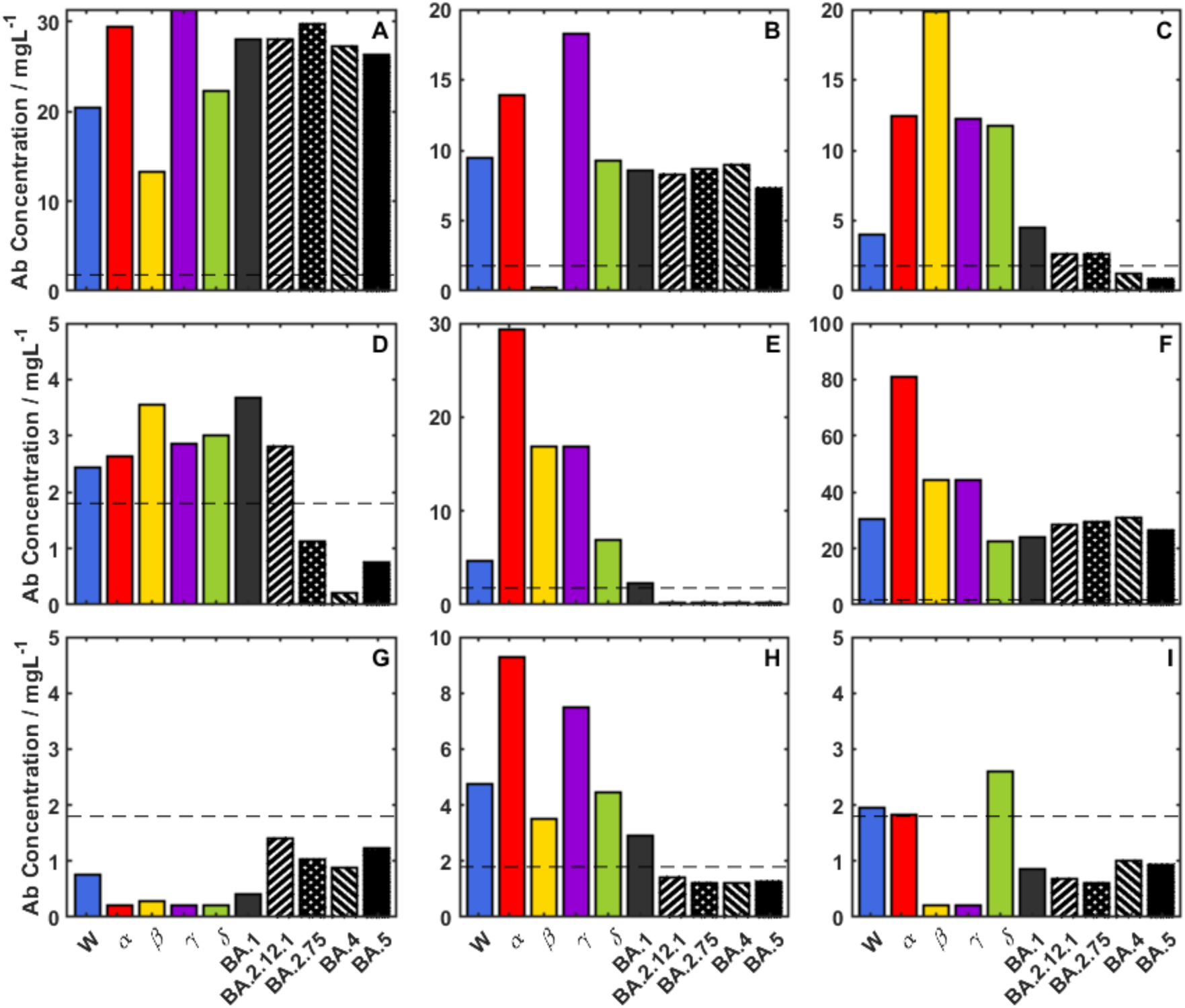
Immunity Endotypes – the dotted line is set at the 1.8 mg/L threshold: (A) a universal positive, U(+) [3× Pfizer vaccination]; (B) Single Dropout β(−) [3× Pfizer vaccination]; (C) Double Dropout BA.4(−) BA.5(−) [[W(+) infection]; (D), Triple Dropout 2.75(−) BA.4(−) BA.5(−) – [2x AZ];(E) Quadruple Dropout BA.2.12(−) BA.2.75(−) BA.4(−) BA.5(−) [1x AZ]; (F) Super Responder [2xAZ - M]; (G) U(−) [2x AZ]; (H) UT() Quadruple dropout that becomes U(+) using half-life rule [W(+)]; (I) U(±) Septuple dropout that changes to quintuple dropout using half-life rule [W(+)] showing higher level of antibody epitope selection.

**Figure 2.**
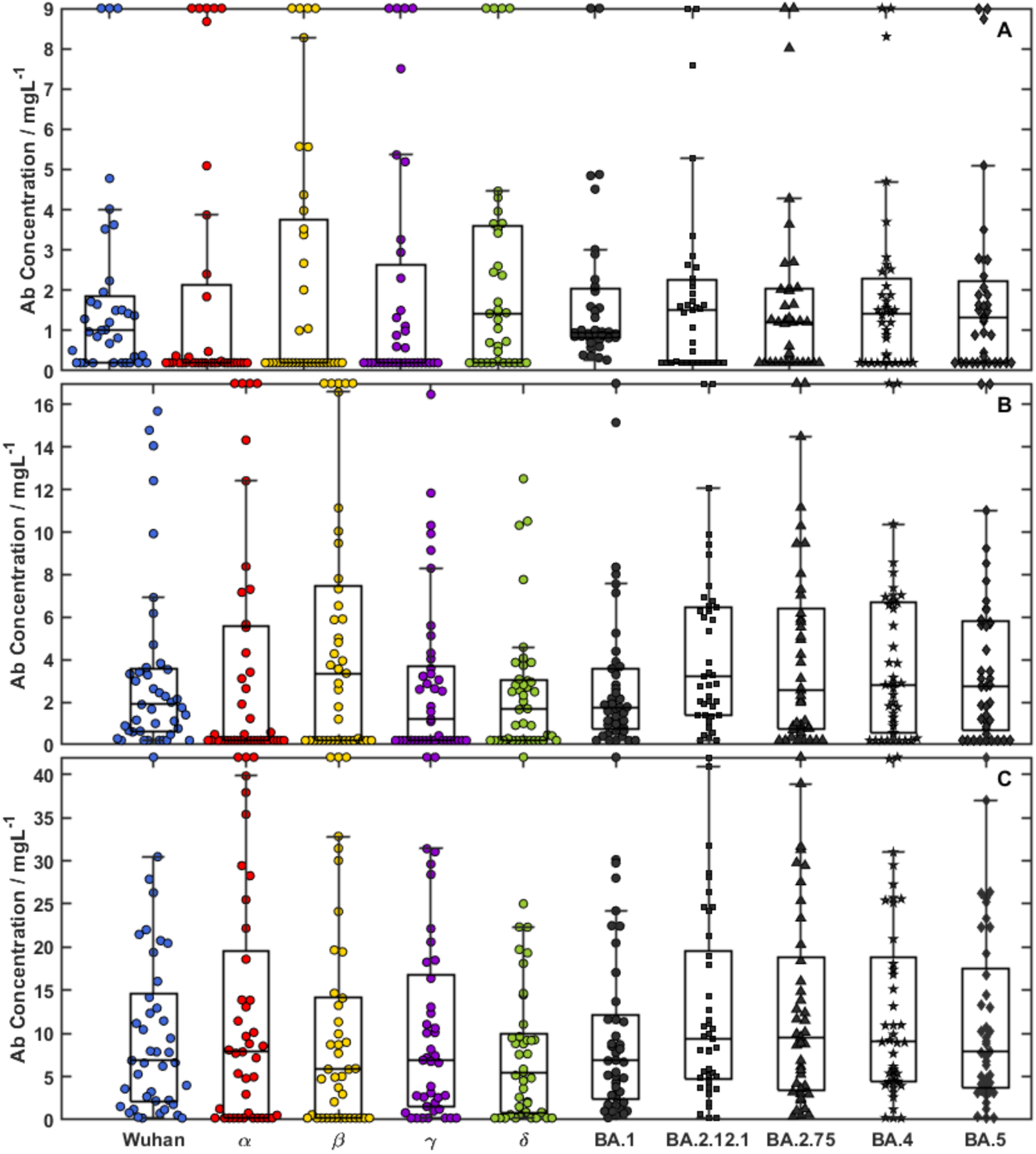
Bee swarm plots for 3 cohorts: (A) W(+); (B) double-vaccinated patients; and (C) triple-vaccinated patients. The boxes show upper and lower quartiles centred on the median and the whiskers indicating the presence of outliers at 1.5× the interquartile range, with the whisker bar falling on the highest/lowest non-outlier data point. Outliers are shown at y-maximum and detailed in Table 1 (Values for the outliers on the top of the plot are shown in Table S7).

The variation in antibody spectra for the W(+) recovery, double-vaccinated and triple-vaccinated cohorts is significant, Figure 1, with the full endotype classification shown in Table 2 (and detailed in Table S8). The ideal antibody response to a universal epitope present in all variant spike proteins, U(+) Figure 1A, was observed in 11% (95% CI 4% - 25%) of the immunologically naïve Wuhan(+) patients, whilst 58% (95% CI 42% - 73%) of patients presented with one or more dropouts, U(±). The most prevalent U(±) was a triple drop-out, Figure 1D, β/γ/δ(−). The initial vaccination cohort has no U(+) endotypes, rising to 22% (95% CI 12% - 37%) in the double-vaccinated cohort and rises again in the triple-vaccinated cohort to 67% (CI 50% - 80%). The double-vaccine groups for AZ or Pfizer offer different protection across the spectrum of variants with AZ showing U((1-8)-) dropouts dominated by U(5-) including the Omicron sub-variants, Figure 1 (F,G,H,I). The Pfizer vaccine, by comparison, only has U(1-4) including a full mix of all variants. In the triple-vaccination cohort, the higher-order drop-outs are nearly eliminated with no U(5) or greater endotypes. One patient with a triple vaccination also had an octuple drop-out, Figure 1I, suggesting a lack of immunity across all variants and immunocompromised.

**Table 2.**
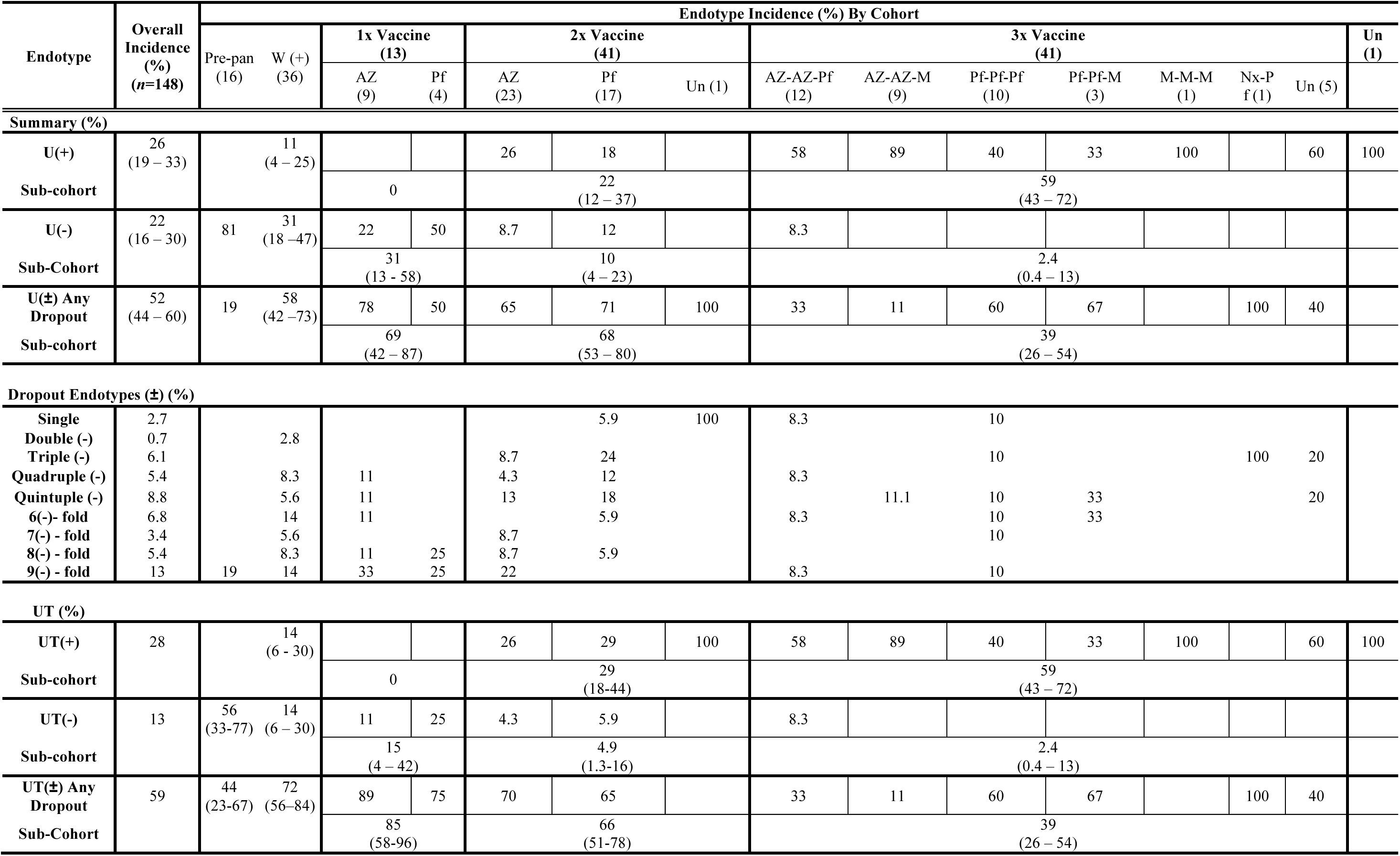
Summary Incidence of the endotype classifications in the 5 cohorts studied. Each incidence is presented as a percentage with the Wilson 95% confidence limits. The individual drop-out classifications are detailed in Table S4.

The variation in classification with time was considered by allowing for the variation of one half-life and doubling all the antibody concentrations to produce a new UT classification. The proportions of each class in the cohorts do not change significantly all falling within the 95% confidence limits.

The cohort distributions of the antibody spectra are shown in Figure 2. The pre-vaccine infection cohort, Figure 2A, shows median antibody levels for Wuhan variants and small levels for Omicron subvariants. These median levels are increased in double and booster vaccination groups, Figure 2(B and C). The interquartile range for Wuhan, natural infection cohort is low, with antibody levels varying from 0.2 – 1.8 mg/L, doubling in the double-vaccinated cohort to 0.6 – 3.6 mg/L and rising further in the triple-boosted cohort, 2 – 14 mg/L. The plots do not show the personalised detail of responses, particularly super- or low-responders; the lower outliers are clustered at the detection limit (200 ng/mL) and the upper outliers are grouped at the top of the figure (Table S7 shows the values for these patients). Of these 17 super-responders, nine show a super-response (beyond the 95^th^ percentile of the population) to more than one variant and three show a full spectrum super-response. The response to one variant can be used to predict the response of others; assuming a linear correlation, the R^2^ (Table S4) and straight-line coefficients are shown in (Table S5)

## Discussion

Four independent patient cohorts were used to explore the pan-variant antibody response to the spike protein of SARS-CoV-2. The natural immune response from naive patients exposed to Wuhan escape variant is compared with single-vaccinated, double-vaccinated and triple-vaccinated cohorts with a control, pre-pandemic cohort. The vaccination cohorts were all targeted at the Wuhan spike protein (first generation vaccines) corresponding to the circulating variant at the time. The antibody response spectra show antibody binding concentrations against the variant spike proteins which would be expected to reflect variant protection against infection and complete recovery. An immunity endotype is defined here using the mechanistic threshold of a clearance and protective serum of 1.8 mg/L (95% CI 0.2–3.4) mg/L measured previously^11^. The threshold is derived from four cohorts: including vaccine response distributions selecting the protection percentiles against live challenge from the Pfizer and AZ vaccines; patients recovering from natural Wuhan infection and circulating NIBSC standards. All measurements were mass-standardised against NIST standard antibody^18^ to produce a fully quantitative result, as were the measurements in this study.

The mechanistic threshold is a variant independent datum level associated with the near-single-variant exposure cohorts and vaccination. The resulting antibody spectra characterise the epitope distributions that are common to variant spike proteins with the caveat that the response is harmonised to the post-translational modification pattern present in the insect cell line (available contemporaneously) from which the spike protein is derived for each variant. The variation of glycosylation patterns^21^ across the variants, the mRNA response of individual patients and infections has significant influence on virus-host interaction, tropism and evasion.

The patient immune response was classified into a number of immune endotypes based on the 1.8 mg/L threshold, U(+), U(−) and U(±). Antibody concentrations wane with a personalised half-life reported to vary between 60 – 200 days for natural antibodies following infection^19^. The collection time of all patient samples is shown in Table 1 is within 60 days of infection or vaccination. The classification was repeated with antibody concentrations doubled to allow for half-life reduction to give UT(+), UT(−) and UT(±). It remains unclear whether variant-specific antibodies can be differentially regulated or whether all clones to SARS-CoV-2 antigens are expressed simultaneously: original antigenic sin^2–4^.

Using the immunity endotype classification, it can be seen there is a significant endotype spectrum of immunity in the patient cohorts, some of which may not be protective against infection even after the booster vaccination. The ideal antibody response produces a universal endotype, U(+), with antibodies binding to conserved endotype on all variants above the protective threshold, Figure 1A. The W(+), pre-vaccination, cohort has antibodies matured against the Wuhan spike protein epitopes during an infection has a U(+) incidence of only 11% (95% CI 4% - 25%). A large proportion did not produce any antibodies above the threshold for all variants, 31 % (95% CI 18% - 47%) with most patients showing at least one gap to a variant in the spectrum, 58% (95% CI 42 % - 73%). The clonal maturation process appears random and certainly not the ‘one and done’ immunity proposition.

The first vaccination cohort did not improve significantly the incidence of the U(+) endotype: 31% (95% CI 13% - 58%) showing a U(−) endotype and 69% (95% CI 42% - 87%) selecting endotypes with more than one drop out – a non-ideal vaccine response but well targeted against the Wuhan variant. The endotype distribution improves with double vaccination and after three doses the U(+) endotype has an incidence of 59% (95% CI 43% - 72%) but the rest of the cohort splitting broadly between U(−) and U(±). Despite population levels of antibodies reported as reasonable, reflecting in the population data in Figure 2, the population immunity for different variant waves, Figure S1, was not well prepared changes in although by the triple vaccination, the cohort had a U(+) incidence of 59% (95% CI 43% - 72%). Repeat exposure to the same mRNA message produced different immune endotype response incidence, breaking original antigenic sin^2–4^.

The original, single vaccination cohort did not produce any U(+) patients indicating that they came from a naïve infection population. However, U(+) incidence in the double and triple vaccination cohorts rise: 22% (95% CI 12% - 37%) and 59% (CI 43% - 72%) respectively, significantly larger. These cohorts are unlikely to be pure having been exposed to the pandemic before, during and after their vaccinations and so reflect a background of natural immunity both from symptomatic and asymptomatic infections. The rising trend however points towards immunity maturation and away from the arguments of original antigenic sin. The mRNA vaccine antibodies may be raised against any part of the spike protein, which is all included in the message, giving rise to a large U(±) endotype presentation, greatest for the double-vaccinated cohort (68% (95% CI 53% – 80%). The U(±) dropout improves significantly with the triple-vaccination cohort falling to 39 % (95% CI 26% – 54%), further pointing to degradation of the original antigenic sin.

The endotypes shown in Figure 1 are snapshots in time all collected with the first half-life of the expected degradation of the antibodies. They are representative of the protection in the population after a booster campaign, but they could be larger by one half-life generating a set endotypes, UT, which have doubled concentrations, Table 2. The result is the number of UT(−) is reduced in the vaccination cohort increasing the UT(±) compared to U((±) from 69% ((95% CI42% – 87%) to 85% (95% CI 58% - 98%), a near-significant change. There is no significant change in the other cohorts suggesting the UT and U endotypes are reasonable representations of immunity profiles in the community. The only other change to the endotype may come with acute infection which antibody levels may rise by 10-fold which may improve the profile of some patients, those close to the limit of detection of the technique. However, it may be concluded that the endotype spectrum is a good estimate of antibody immunity.

Structure function and epitope stability are essential to optimising vaccines against a target that is mutating rapidly such as the significant mutations to the Omicron subvariants occurring in 70 days^22^ with 100-day vaccine responses^23^. A hemispherical region at the top of the spike S1 region, Figure 3, is relevant to antibody protection^24^ via both neutralisation and opsonisation. In addition, a second epitope region around the hinge region would prevent the pre-fusion-fusion transition. By contrast, the receptor binding domain (RBD) binding region has significant build-up of mutations affecting infectivity and immune-evasiveness of each strain: a kinetic epitope mapping study^25^ based on clones from the universal endotype could be useful in the design of a universal ‘Hinge Vaccine’ and new monoclonals for immunotherapy.

**Figure 3.**
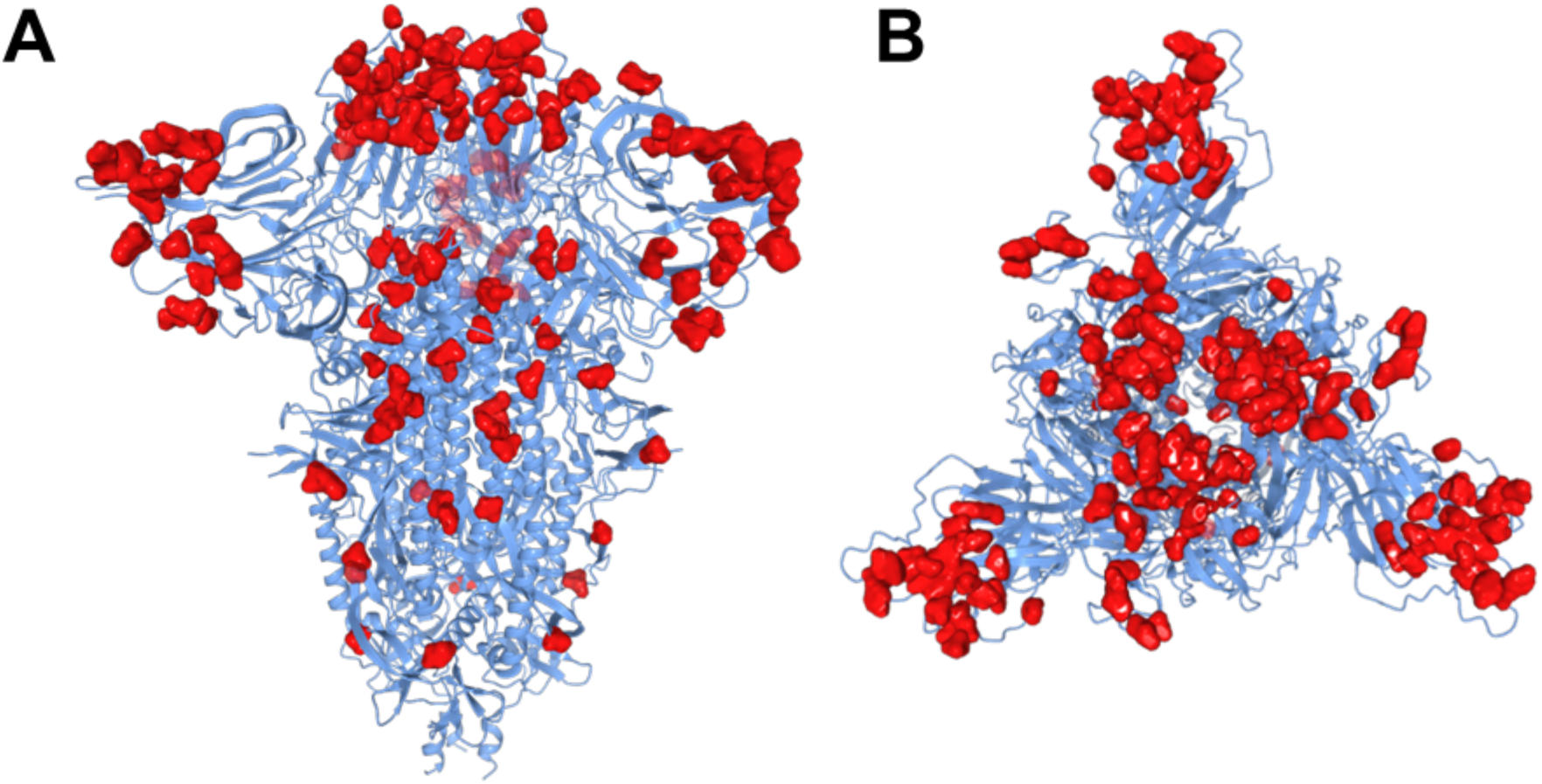
The superimposed Alpha, Beta, Gamma, Delta, BA.1, BA.2.12.1, BA.4/5 mutations of the SARS-CoV-2 Spike protein shown from A) side-on and B) top-down views. Image created using the Swiss Institute of Bioinformatics Expasy tool^26^.

## Conclusions

A set of mechanistic thresholds or serum characteristics that are variant bridging makes the assessment of infection and vaccine efficacy comparable between cohorts. The mass-standardised variant immunity spectrum reveals a set of immunity endotypes in each of the cohorts with a rising incidence of U(+) the universal conserved endotype response. The host response is optimised to epitopes providing recovery, producing memory B cells and T cells with chosen epitope regions of the proteins, potentially unique to each host and leading to immunity imprinting^27–29^ and original antigenic sin^2–4^. The immunity imprint is not conserved in three cohorts studied here with the number of drop-out endotypes falling with vaccination and infection. Indeed, the evolution away from drop-out endotypes towards the U(+) universal response points to a universal vaccine or immunotherapy and a route to suppression of the virus. However, there are 39% of patients with U(±) and worse up to 10% with U(−), for which a good quantity of antibodies to the infecting variant is never present. The low concentration and potentially low affinity, define a non-sterilising serum leaving patients not able to clear the virus and vulnerable to post-viral sequalae consistent with the persistent virus hypothesis for long covid^30–33^.

## Data Availability

All data produced in the present study are available upon reasonable request to the authors

## Acknowledgements

The authors would like to thank Dr Jonathan Snicker for their guidance on the potential strategic and policy implications of the scientific findings.

## Supplementary Data

**Table S1.**
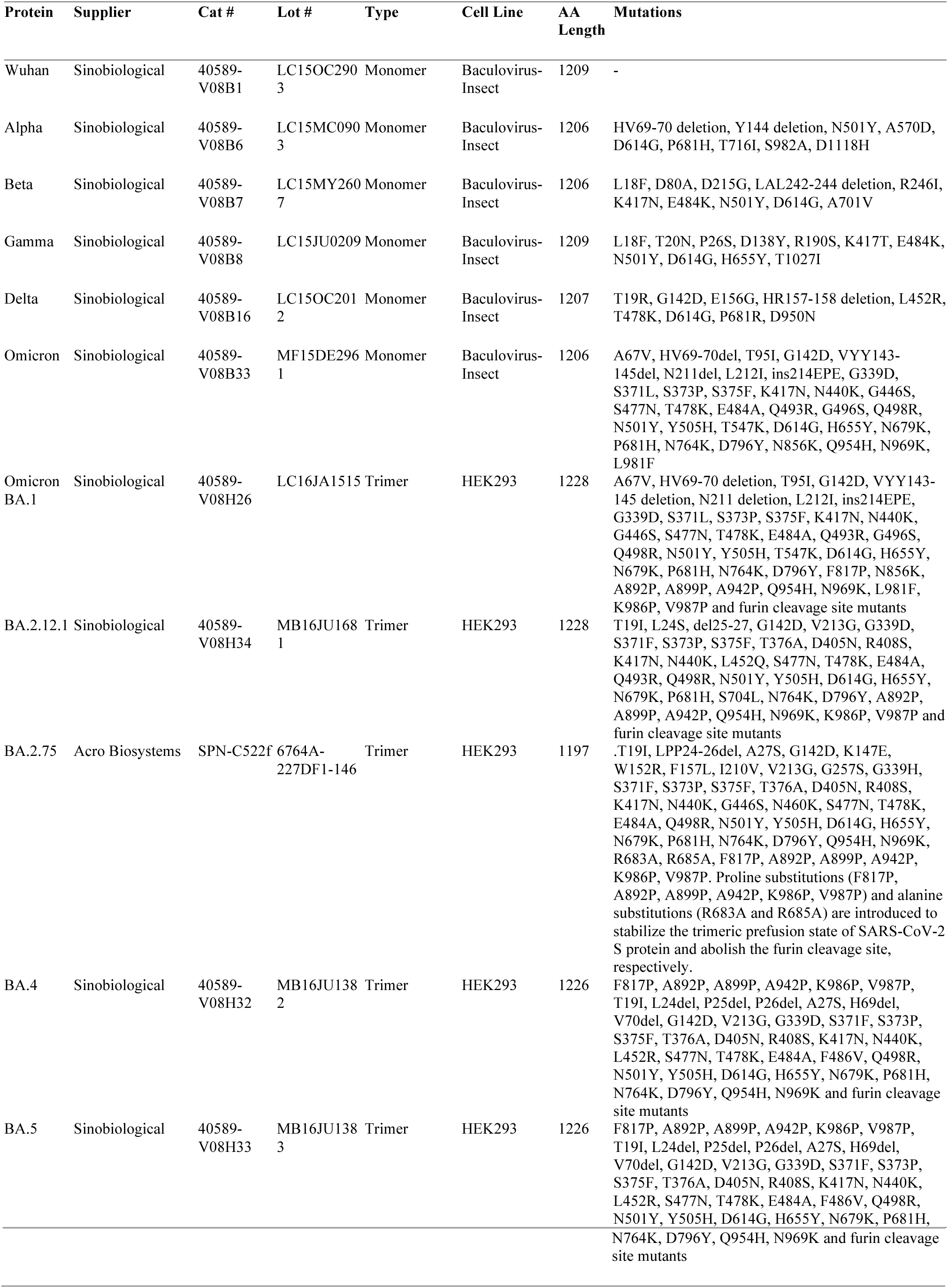
Variant Spike proteins, manufacturer, cell line, modifications, length and mutations.

**Table S2.**
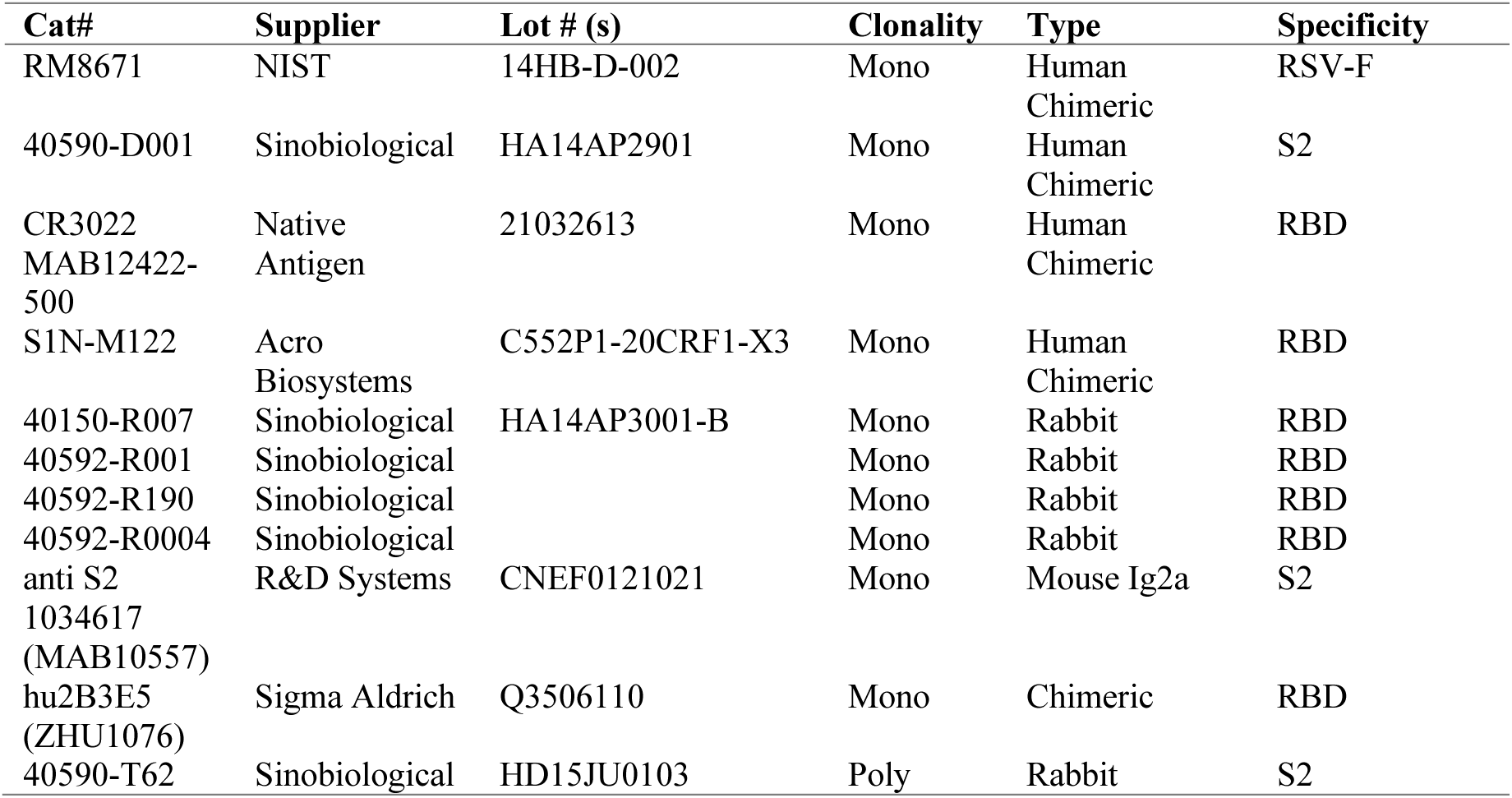
Panel of Antibodies used to screen Spike protein integrity on the surface.

**Table S3.**
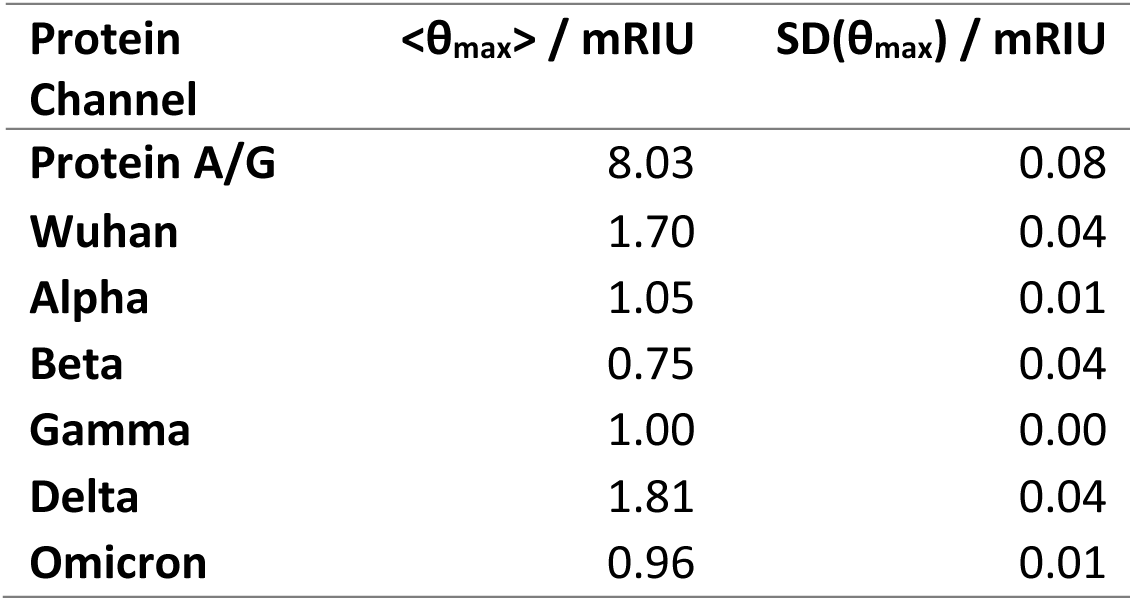
Mean and Standard Deviation of θmax values determined from the binding of aS2 antibody 40590-D001 to each protein channel. As this is the first time out – shall we include this??

**Figure S1.**
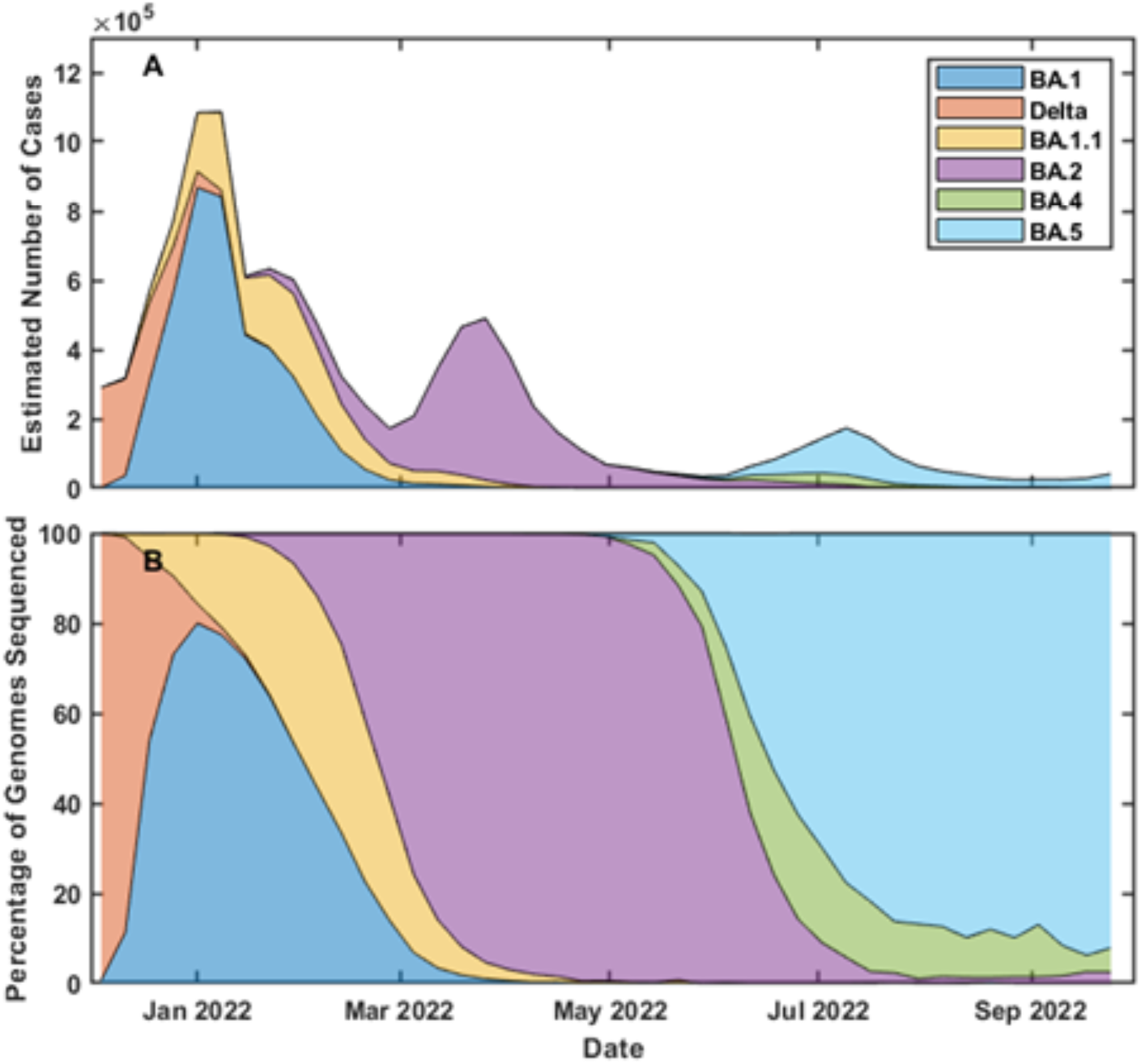
Prevalent SARS-CoV-2 variants in the UK November 2021-September 2022. A) Estimated number of cases and B) Variant share as % of genomes sequenced from a random sample. Data is provided by the Wellcome Sanger Institut ^34^

**Table S4.** Table of Line-of-best fit gradients, column = row × gradient +intercept. The intercept is the Limit of Detection.

| Wuhan (+) | $\alpha$ | $\beta$ | $\gamma$ | $\delta$ | Wuhan | BA.1 | BA.2.12.1 | BA.2.75 | BA.4 | BA.5 |
| --- | --- | --- | --- | --- | --- | --- | --- | --- | --- | --- |
| $\alpha$ | 1.00 | 0.42 | 0.39 | 0.30 | 0.21 | 0.07 | 0.05 | 0.04 | 0.06 | 0.05 |
| $\beta$ | 2.30 | 1.00 | 0.91 | 0.71 | 0.50 | 0.17 | 0.12 | 0.10 | 0.13 | 0.12 |
| $\gamma$ | 2.51 | 1.08 | 1.00 | 0.79 | 0.56 | 0.19 | 0.15 | 0.12 | 0.16 | 0.15 |
| $\delta$ | 3.04 | 1.31 | 1.23 | 1.00 | 0.73 | 0.26 | 0.23 | 0.19 | 0.24 | 0.23 |
| Wuhan | 3.98 | 1.72 | 1.64 | 1.35 | 1.00 | 0.37 | 0.35 | 0.28 | 0.35 | 0.35 |
| BA.1 | 9.09 | 4.02 | 3.89 | 3.32 | 2.51 | 1.00 | 1.05 | 0.86 | 1.01 | 1.04 |
| BA.2.12.1 | 4.52 | 2.06 | 2.16 | 2.09 | 1.70 | 0.75 | 1.00 | 0.83 | 0.91 | 0.98 |
| BA.2.75 | 5.07 | 2.32 | 2.47 | 2.42 | 1.99 | 0.88 | 1.19 | 1.00 | 1.09 | 1.17 |
| BA.4 | 5.93 | 2.62 | 2.70 | 2.51 | 2.01 | 0.85 | 1.07 | 0.89 | 1.00 | 1.06 |
| BA.5 | 4.90 | 2.19 | 2.30 | 2.20 | 1.78 | 0.77 | 1.02 | 0.85 | 0.94 | 1.00 |
| **Double Vaccinated** |  |  |  |  |  |  |  |  |  |  |
| $\alpha$ | 1.00 | 0.64 | 0.14 | 0.14 | 0.24 | 0.14 | 0.11 | 0.13 | 0.11 | 0.11 |
| $\beta$ | 1.11 | 1.00 | 0.18 | 0.13 | 0.25 | 0.13 | 0.06 | 0.08 | 0.06 | 0.07 |
| $\gamma$ | 2.03 | 1.42 | 1.00 | 0.67 | 0.76 | 0.79 | 0.90 | 0.90 | 0.94 | 0.89 |
| $\delta$ | 3.30 | 1.83 | 1.12 | 1.00 | 1.26 | 1.15 | 1.38 | 1.40 | 1.39 | 1.32 |
| Wuhan | 2.87 | 1.74 | 0.65 | 0.64 | 1.00 | 0.81 | 1.00 | 1.07 | 1.01 | 0.97 |
| BA.1 | 1.72 | 0.94 | 0.71 | 0.61 | 0.85 | 1.00 | 1.38 | 1.38 | 1.35 | 1.30 |
| BA.2.12.1 | 0.61 | 0.20 | 0.36 | 0.33 | 0.47 | 0.62 | 1.00 | 1.00 | 0.98 | 0.94 |
| BA.2.75 | 0.72 | 0.26 | 0.34 | 0.32 | 0.48 | 0.59 | 0.95 | 1.00 | 0.95 | 0.92 |
| BA.4 | 0.62 | 0.19 | 0.37 | 0.33 | 0.47 | 0.60 | 0.98 | 1.00 | 1.00 | 0.95 |
| BA.5 | 0.68 | 0.25 | 0.39 | 0.35 | 0.50 | 0.64 | 1.03 | 1.06 | 1.04 | 1.00 |
| **Triple Vaccinated** |  |  |  |  |  |  |  |  |  |  |
| $\alpha$ | 1.00 | 0.57 | 0.67 | 0.45 | 0.50 | 0.49 | 0.48 | 0.48 | 0.46 | 0.45 |
| $\beta$ | 1.11 | 1.00 | 0.79 | 0.56 | 0.61 | 0.59 | 0.56 | 0.56 | 0.53 | 0.52 |
| $\gamma$ | 1.27 | 0.77 | 1.00 | 0.73 | 0.79 | 0.83 | 0.81 | 0.80 | 0.76 | 0.75 |
| $\delta$ | 1.46 | 0.91 | 1.24 | 1.00 | 1.08 | 1.16 | 1.15 | 1.12 | 1.06 | 1.06 |
| Wuhan | 1.32 | 0.82 | 1.10 | 0.89 | 1.00 | 1.04 | 1.06 | 1.05 | 1.00 | 0.98 |
| BA.1 | 1.14 | 0.69 | 1.00 | 0.83 | 0.91 | 1.00 | 1.02 | 0.99 | 0.94 | 0.94 |
| BA.2.12.1 | 1.00 | 0.59 | 0.89 | 0.75 | 0.84 | 0.92 | 1.00 | 0.97 | 0.94 | 0.92 |
| BA.2.75 | 1.02 | 0.61 | 0.90 | 0.75 | 0.85 | 0.92 | 1.00 | 1.00 | 0.96 | 0.92 |
| BA.4 | 1.06 | 0.62 | 0.92 | 0.76 | 0.86 | 0.94 | 1.04 | 1.03 | 1.00 | 0.96 |
| BA.5 | 1.09 | 0.65 | 0.97 | 0.81 | 0.91 | 1.00 | 1.08 | 1.06 | 1.03 | 1.00 |

**Table S5.** Table of R^2^ values.

| Wuhan (+) | $\alpha$ | $\beta$ | $\gamma$ | $\delta$ | Wuhan | BA.1 | BA.2.12.1 | BA.2.75 | BA.4 | BA.5 |
| --- | --- | --- | --- | --- | --- | --- | --- | --- | --- | --- |
| $\alpha$ | 1.00 | 0.99 | 0.99 | 0.96 | 0.92 | 0.80 | 0.47 | 0.44 | 0.57 | 0.50 |
| $\beta$ | 0.99 | 1.00 | 0.99 | 0.96 | 0.92 | 0.83 | 0.50 | 0.47 | 0.59 | 0.52 |
| $\gamma$ | 0.99 | 0.99 | 1.00 | 0.98 | 0.96 | 0.87 | 0.57 | 0.55 | 0.66 | 0.60 |
| $\delta$ | 0.96 | 0.96 | 0.98 | 1.00 | 0.99 | 0.93 | 0.69 | 0.67 | 0.77 | 0.71 |
| Wuhan | 0.92 | 0.92 | 0.96 | 0.99 | 1.00 | 0.96 | 0.77 | 0.75 | 0.84 | 0.79 |
| BA.1 | 0.80 | 0.83 | 0.87 | 0.93 | 0.96 | 1.00 | 0.89 | 0.87 | 0.92 | 0.89 |
| BA.2.12.1 | 0.47 | 0.50 | 0.57 | 0.69 | 0.77 | 0.89 | 1.00 | 1.00 | 0.99 | 1.00 |
| BA.2.75 | 0.44 | 0.47 | 0.55 | 0.67 | 0.75 | 0.87 | 1.00 | 1.00 | 0.99 | 0.99 |
| BA.4 | 0.57 | 0.59 | 0.66 | 0.77 | 0.84 | 0.92 | 0.99 | 0.99 | 1.00 | 1.00 |
| BA.5 | 0.50 | 0.52 | 0.60 | 0.71 | 0.79 | 0.89 | 1.00 | 0.99 | 1.00 | 1.00 |
| **Double Vaccinated** |  |  |  |  |  |  |  |  |  |  |
| $\alpha$ | 1.00 | 0.84 | 0.54 | 0.68 | 0.82 | 0.48 | 0.26 | 0.31 | 0.26 | 0.27 |
| $\beta$ | 0.84 | 1.00 | 0.50 | 0.50 | 0.66 | 0.35 | 0.11 | 0.15 | 0.11 | 0.13 |
| $\gamma$ | 0.54 | 0.50 | 1.00 | 0.86 | 0.70 | 0.75 | 0.57 | 0.56 | 0.59 | 0.59 |
| $\delta$ | 0.68 | 0.50 | 0.86 | 1.00 | 0.90 | 0.84 | 0.67 | 0.67 | 0.68 | 0.68 |
| Wuhan | 0.82 | 0.66 | 0.70 | 0.90 | 1.00 | 0.83 | 0.68 | 0.72 | 0.69 | 0.69 |
| BA.1 | 0.48 | 0.35 | 0.75 | 0.84 | 0.83 | 1.00 | 0.92 | 0.90 | 0.90 | 0.91 |
| BA.2.12.1 | 0.26 | 0.11 | 0.57 | 0.67 | 0.68 | 0.92 | 1.00 | 0.97 | 0.98 | 0.98 |
| BA.2.75 | 0.31 | 0.15 | 0.56 | 0.67 | 0.72 | 0.90 | 0.97 | 1.00 | 0.98 | 0.98 |
| BA.4 | 0.26 | 0.11 | 0.59 | 0.68 | 0.69 | 0.90 | 0.98 | 0.98 | 1.00 | 1.00 |
| BA.5 | 0.27 | 0.13 | 0.59 | 0.68 | 0.69 | 0.91 | 0.98 | 0.98 | 1.00 | 1.00 |
| **Triple Vaccinated** |  |  |  |  |  |  |  |  |  |  |
| $\alpha$ | 1.00 | 0.79 | 0.92 | 0.81 | 0.81 | 0.75 | 0.69 | 0.70 | 0.70 | 0.70 |
| $\beta$ | 0.79 | 1.00 | 0.78 | 0.71 | 0.71 | 0.64 | 0.57 | 0.58 | 0.57 | 0.58 |
| $\gamma$ | 0.92 | 0.78 | 1.00 | 0.96 | 0.94 | 0.91 | 0.85 | 0.85 | 0.84 | 0.86 |
| $\delta$ | 0.81 | 0.71 | 0.96 | 1.00 | 0.98 | 0.98 | 0.93 | 0.92 | 0.90 | 0.93 |
| Wuhan | 0.81 | 0.71 | 0.94 | 0.98 | 1.00 | 0.97 | 0.94 | 0.95 | 0.93 | 0.94 |
| BA.1 | 0.75 | 0.64 | 0.91 | 0.98 | 0.97 | 1.00 | 0.97 | 0.96 | 0.94 | 0.97 |
| BA.2.12.1 | 0.69 | 0.57 | 0.85 | 0.93 | 0.94 | 0.97 | 1.00 | 0.98 | 0.99 | 1.00 |
| BA.2.75 | 0.70 | 0.58 | 0.85 | 0.92 | 0.95 | 0.96 | 0.98 | 1.00 | 0.99 | 0.99 |
| BA.4 | 0.70 | 0.57 | 0.84 | 0.90 | 0.93 | 0.94 | 0.99 | 0.99 | 1.00 | 0.99 |
| BA.5 | 0.70 | 0.58 | 0.86 | 0.93 | 0.94 | 0.97 | 1.00 | 0.99 | 0.99 | 1.00 |

**Table S6.** Table of medians and quartiles, broken down by exposure cohort. All values are in mg L^-1^.

| **Pre-Pandemic** | **$\alpha$** | **$\beta$** | **$\gamma$** | **$\delta$** | **Wuhan** | **BA.1** | **BA.2.12.1** | **BA.2.75** | **BA.4** | **BA.5** |
| --- | --- | --- | --- | --- | --- | --- | --- | --- | --- | --- |
| Lower Quartile | 0.2 | 0.2 | 0.2 | 0.2 | 0.2 | 0.2 | 0.2 | 0.2 | 0.2 | 0.2 |
| Median | 0.2 | 0.2 | 0.2 | 0.2 | 0.2 | 0.4 | 0.2 | 0.2 | 0.3 | 0.3 |
| Upper Quartile | 0.6 | 0.2 | 0.2 | 0.2 | 0.2 | 0.7 | 1.0 | 0.7 | 0.9 | 0.9 |
| **Wuhan (+)** |  |  |  |  |  |  |  |  |  |  |
| Lower Quartile | 0.2 | 0.2 | 0.2 | 0.2 | 0.2 | 0.8 | 0.2 | 0.2 | 0.2 | 0.2 |
| Median | 0.2 | 0.2 | 0.2 | 1.4 | 1.0 | 1.0 | 1.5 | 1.2 | 1.4 | 1.3 |
| Upper Quartile | 2.1 | 3.8 | 2.6 | 3.6 | 1.8 | 2.0 | 2.2 | 2.0 | 2.3 | 2.2 |
| **Double Vaccinated** |  |  |  |  |  |  |  |  |  |  |
| Lower Quartile | 0.2 | 0.2 | 0.2 | 0.2 | 0.6 | 0.8 | 1.4 | 0.7 | 0.5 | 0.7 |
| Median | 0.2 | 3.4 | 1.2 | 1.7 | 1.9 | 1.8 | 3.2 | 2.6 | 2.8 | 2.8 |
| Upper Quartile | 5.6 | 7.5 | 3.7 | 3.0 | 3.6 | 3.6 | 6.5 | 6.4 | 6.7 | 5.8 |
| **Triple Vaccinated** |  |  |  |  |  |  |  |  |  |  |
| Lower Quartile | 0.2 | 0.2 | 1.5 | 0.8 | 2.1 | 2.4 | 4.8 | 3.5 | 4.4 | 3.7 |
| Median | 7.9 | 5.8 | 6.9 | 5.5 | 6.9 | 6.8 | 9.4 | 9.6 | 9.0 | 7.9 |
| Upper Quartile | 19.5 | 14.2 | 16.9 | 10.0 | 14.6 | 12.1 | 19.5 | 18.9 | 18.8 | 17.4 |

**Table S7.**
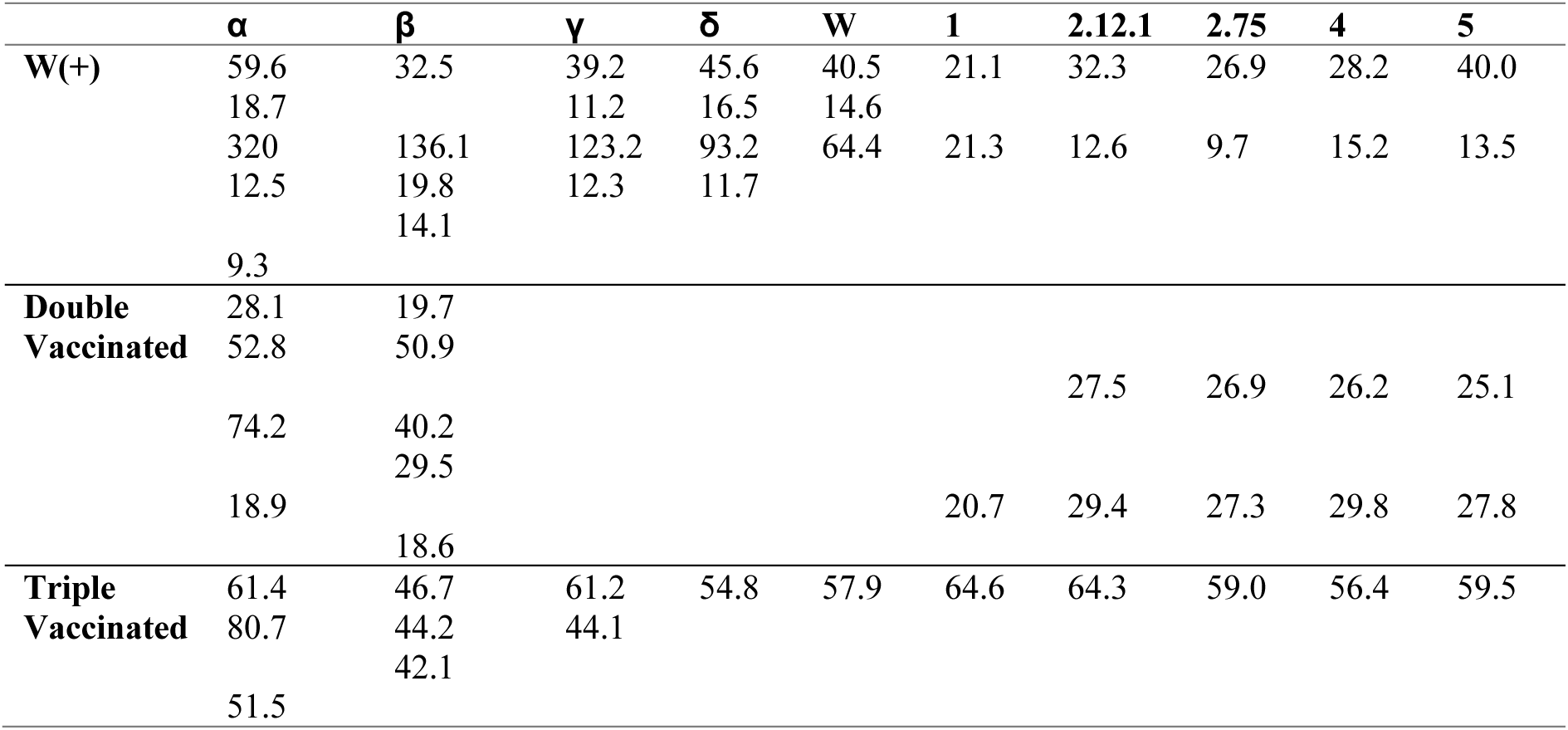
Antibody concentrations for the Patients present in Figure 3 displayed on the upper limit of the figure.

**Table S8.**
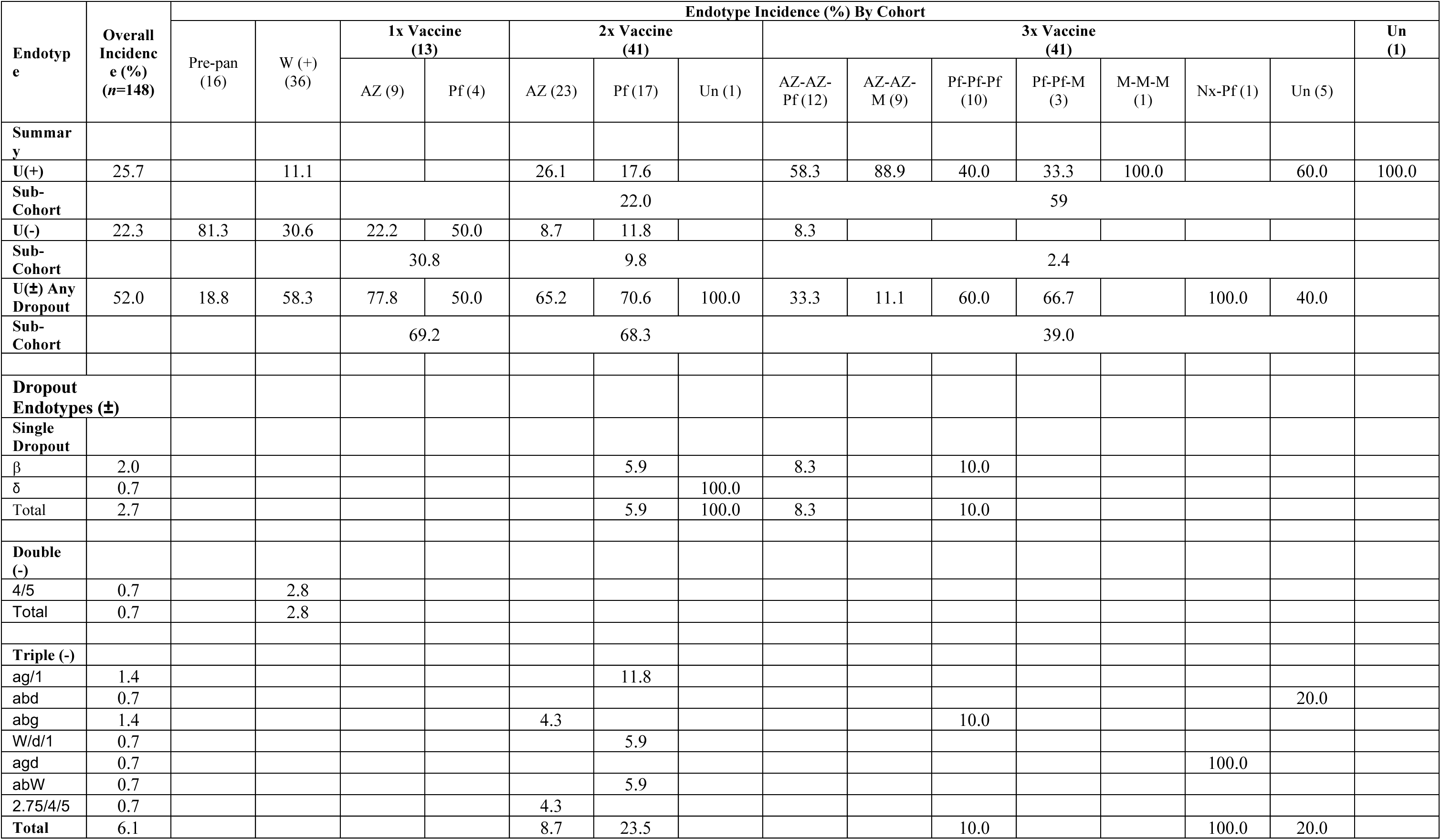
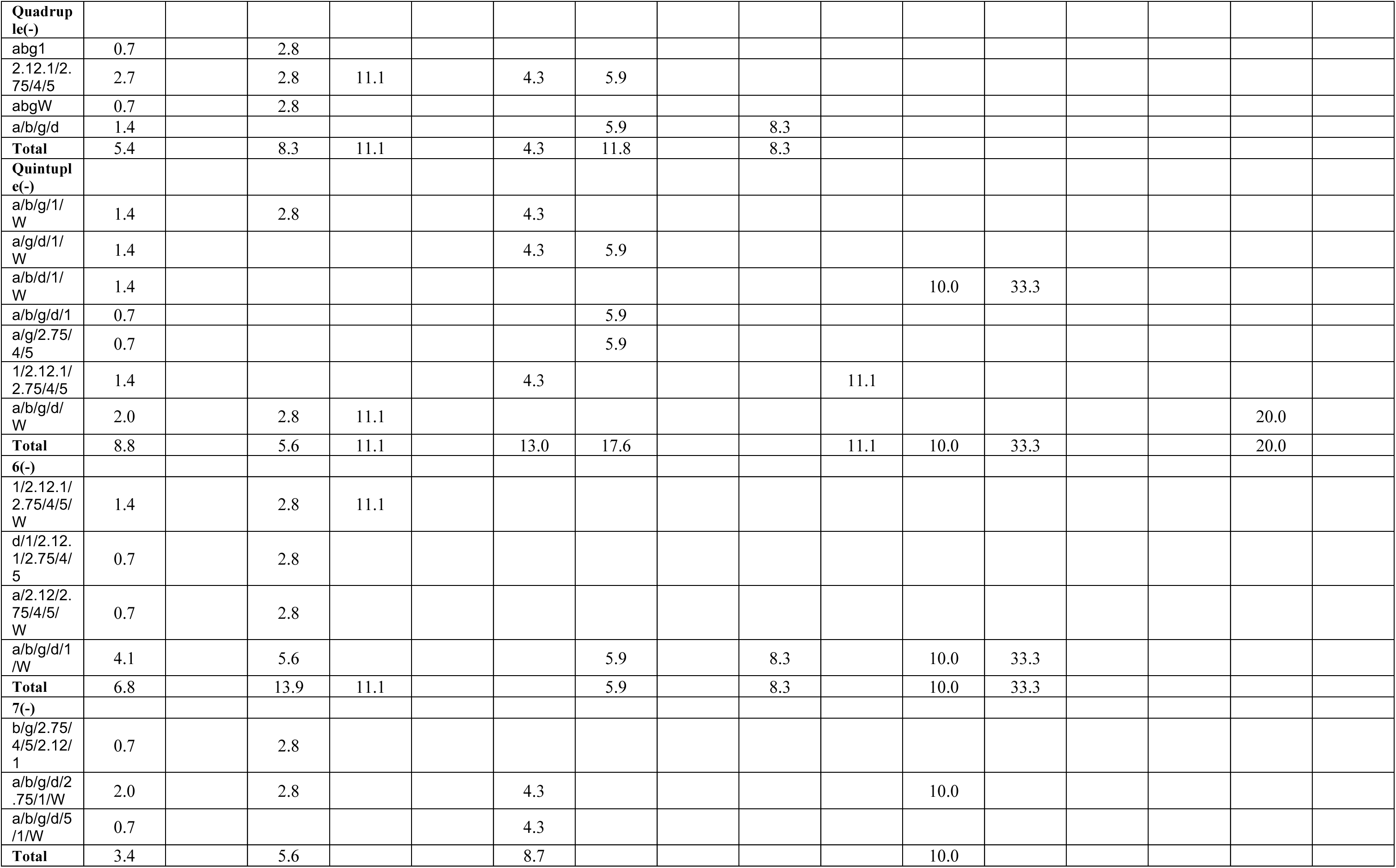
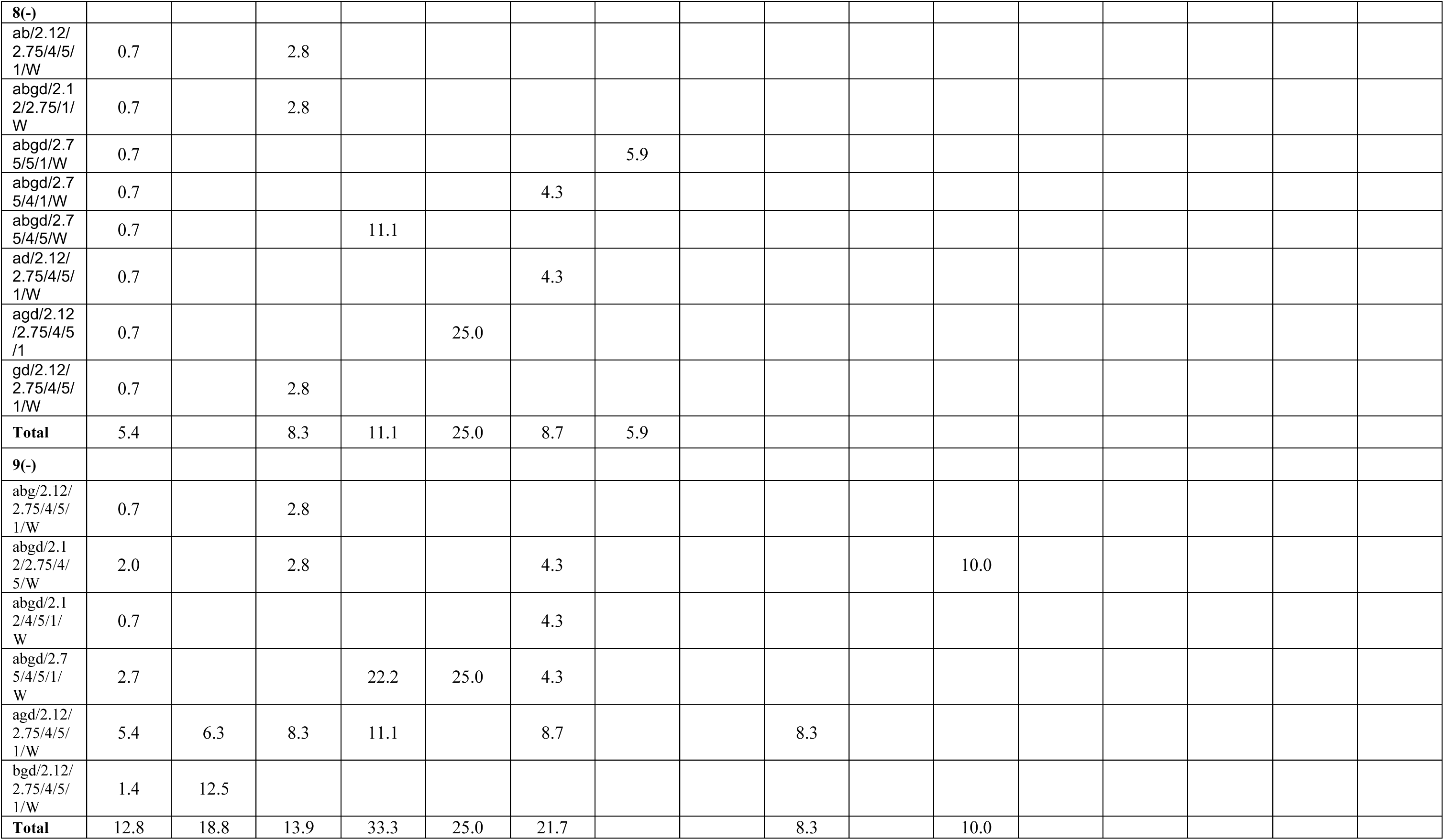
Full Endotype Profile for the cohort detailing the dropout variants.

## References

1. MacLean AJ, Deimel LP, Zhou P, et al. Affinity maturation of antibody responses is mediated by differential plasma cell proliferation. Science 2025; 387(6732): 413–20.

2. Evans JP, Liu SL. Challenges and Prospects in Developing Future SARS-CoV-2 Vaccines: Overcoming Original Antigenic Sin and Inducing Broadly Neutralizing Antibodies. J Immunol 2023; 211(10): 1459–67.

3. Spatola M, Nziza N, Jung W, et al. Neurologic sequelae of COVID-19 are determined by immunologic imprinting from previous coronaviruses. Brain 2023; 146(10): 4292–305.

4. Reynolds CJ, Pade C, Gibbons JM, et al. Immune boosting by B.1.1.529 (Omicron) depends on previous SARS-CoV-2 exposure. Science 2022: eabq1841.

5. Markov PV, Ghafari M, Beer M, et al. The evolution of SARS-CoV-2. Nature Reviews Microbiology 2023; 21(6): 361–79.

6. Scheaffer SM, Lee D, Whitener B, et al. Bivalent SARS-CoV-2 mRNA vaccines increase breadth of neutralization and protect against the BA.5 Omicron variant in mice. Nature Medicine 2022.

7. Chalkias S, Harper C, Vrbicky K, et al. A Bivalent Omicron-Containing Booster Vaccine against Covid-19. The New England journal of medicine 2022; 387(14): 1279–91.

8. Pérez-Then E, Lucas C, Monteiro VS, et al. Neutralizing antibodies against the SARS-CoV-2 Delta and Omicron variants following heterologous CoronaVac plus BNT162b2 booster vaccination. Nat Med 2022; 28(3): 481–5.

9. Ai J, Zhang H, Zhang Y, et al. Omicron variant showed lower neutralizing sensitivity than other SARS-CoV-2 variants to immune sera elicited by vaccines after boost. Emerg Microbes Infect 2022; 11(1): 337–43.

10. King DF, Groves H, Weller C, et al. Realising the potential of correlates of protection for vaccine development, licensure and use: short summary. npj Vaccines 2024; 9(1): 82.

11. James-Pemberton PH, Helliwell MW, Olkhov RV, et al. Mass-Standardised Quantitative Measurements of the Antibody Levels for SARS-CoV-2 beyond Correlates of Protection and Clearance medRxiv 2025: 2022.07.12.22277533.

12. Sun K, Bhiman JN, Tempia S, et al. SARS-CoV-2 correlates of protection from infection against variants of concern. Nat Med 2024; 30(10): 2805–12.

13. Han P, Li L, Liu S, et al. Receptor binding and complex structures of human ACE2 to spike RBD from omicron and delta SARS-CoV-2. Cell 2022; 185(4): 630–40.e10.

14. Barton MI, MacGowan SA, Kutuzov MA, Dushek O, Barton GJ, van der Merwe PA. Effects of common mutations in the SARS-CoV-2 Spike RBD and its ligand, the human ACE2 receptor on binding affinity and kinetics. eLife 2021; 10: e70658.

15. Granerud BK, Ueland T, Lind A, et al. Omicron Variant Generates a Higher and More Sustained Viral Load in Nasopharynx and Saliva Than the Delta Variant of SARS-CoV-2. Viruses 2022; 14(11).

16. Muecksch F, Wise H, Templeton K, et al. Longitudinal variation in SARS-CoV-2 antibody levels and emergence of viral variants: a serological analysis. Lancet Microbe 2022; 3(7): e493–e502.

17. Shaw AM, Hyde C, Merrick B, et al. Real-world evaluation of a novel technology for quantitative simultaneous antibody detection against multiple SARS-CoV-2 antigens in a cohort of patients presenting with COVID-19 syndrome. Analyst 2020.

18. Formolo T LM, Levy M, Kilpatrick L, Lute S, Phinney K, et al.. Determination of the NISTmAb primary structure. State-of-the-Art and Emerging Technologies for Therapeutic Monoclonal Antibody Characterization: ACS Symposium Series; 2015: 1–62.

19. Dan JM, Mateus J, Kato Y, et al. Immunological memory to SARS-CoV-2 assessed for up to 8 months after infection. Science 2021; 371(6529): eabf4063.

20. McLellan JS, Chen M, Kim A, Yang Y, Graham BS, Kwong PD. Structural basis of respiratory syncytial virus neutralization by motavizumab. Nature structural & molecular biology 2010; 17(2): 248–50.

21. Onigbinde S, Reyes CDG, Fowowe M, et al. Variations in O-Glycosylation Patterns Influence Viral Pathogenicity, Infectivity, and Transmissibility in SARS-CoV-2 Variants. Biomolecules 2023; 13(10): 1467.

22. Institute S. 2022. https://covid19.sanger.ac.uk/lineages/raw (accessed September 2022.

23. Pfizer. Pfizer and BioNTech Provide Update on Omicron Variant. 2021.

24. Harvey WT, Carabelli AM, Jackson B, et al. SARS-CoV-2 variants, spike mutations and immune escape. Nature Reviews Microbiology 2021; 19(7): 409–24.

25. Read T, Olkhov RV, Williamson ED, Shaw AM. Kinetic epitope mapping of monoclonal antibodies raised against the Yersinia pestis virulence factor LcrV. Biosensors & bioelectronics 2015; 65: 47–53.

26. ViralZone SSIoB. Spike Protein Figure. 2022. https://viralzone.expasy.org/9556 (accessed 21 June 2022).

27. Röltgen K, Nielsen SCA, Silva O, et al. Immune imprinting, breadth of variant recognition, and germinal center response in human SARS-CoV-2 infection and vaccination. Cell 2022; 185(6): 1025–40.e14.

28. Wheatley AK, Fox A, Tan HX, et al. Immune imprinting and SARS-CoV-2 vaccine design. Trends Immunol 2021; 42(11): 956–9.

29. Reynolds CJ, Pade C, Gibbons JM, et al. Immune boosting by B.1.1.529 (Omicron) depends on previous SARS-CoV-2 exposure. Science 2022; 377(6603): eabq1841.

30. Altmann DM, Whettlock EM, Liu S, Arachchillage DJ, Boyton RJ. The immunology of long COVID. Nature Reviews Immunology 2023; 23(10): 618–34.

31. Yong SJ. Long COVID or post-COVID-19 syndrome: putative pathophysiology, risk factors, and treatments. Infectious Diseases 2021; 53(10): 737–54.

32. Davis HE, McCorkell L, Vogel JM, Topol EJ. Long COVID: major findings, mechanisms and recommendations. Nature Reviews Microbiology 2023; 21(3): 133–46.

33. Klein J, Wood J, Jaycox JR, et al. Distinguishing features of long COVID identified through immune profiling. Nature 2023; 623(7985): 139–48.

34. COVID-19 Genomic Surveillance. 2022. https://covid19.sanger.ac.uk/lineages/raw?latitude=51.622741&longitude=-1.455710&zoom=4.708800&p_type=area&cases_type=area&xMin=2021-12-04&xMax=2022-09-24 (accessed 24/09/2022 2022).

